# Dexmedetomidine as a Perioperative Adjunct in Valvular Cardiac Surgery

**DOI:** 10.1101/2025.09.04.25334916

**Authors:** Jalaram Harshappan, Sanjeeta Umbarkar, Parimal Pimpalkhute, Pooja Pimpalkhute, Renu Upadhyay, Juhi Kacha

## Abstract

**Background:** Valvular heart surgery is associated with considerable perioperative risks including hemodynamic instability, arrhythmias, renal dysfunction and prolonged ICU stay. Dexmedetomidine a selective α2-adrenoceptor agonist, provides sedation and sympatholysis and has been investigated as a cardioprotective adjunct.

**Objectives:** To evaluate the effects of dexmedetomidine on intraoperative hemodynamic stability and postoperative outcomes in adult patients undergoing valvular replacement surgery.

**Methods:** In this retrospective study, 154 patients who underwent valvular surgery under cardiopulmonary bypass between January 2022 and June 2023 were analyzed. Patients were divided into Group C (control, n=77) and Group D (dexmedetomidine, n=77). Group D received a continuous infusion of dexmedetomidine (0.5 μg/kg/hr) from induction until the end of surgery. Hemodynamic and depth of sedation parameters (HR, MAP, CVP, NIRS, entropy) were measured at predefined intraoperative time points. Postoperative outcomes included time of extubation, atrial fibrillation (AF), acute kidney injury (AKI), Type 1 neurological injury, reintubation, reoperation, ICU readmission, ICU length of stay (LOS), and 30-day mortality.

**Results:** Baseline demographics were similar between groups. Group D exhibited significantly lower HR at post-induction, sternotomy, and pericardiotomy compared with Group C (p<0.001 at all three time points). MAP was comparable between groups at post-induction, sternotomy, and pericardiotomy, although Group D showed a significantly higher MAP at 5 minutes after protamine administration (77.36 ± 7.66 vs. 73.56 ± 8.16 mmHg, p=0.003). ICU LOS was numerically shorter in Group D but did not reach statistical significance (8.19 ± 4.51 vs. 8.68 ± 3.88 days, p=0.480). Time to extubation was also numerically lower in Group D but was not statistically significant (173.04 ± 377.63 vs. 192.10 ± 352.61 hours, p=0.747). Postoperative complications were numerically lower in Group D, including AF (11.7% vs. 13.0%, p=1.000), AKI (11.7% vs. 14.3%, p=0.811), reintubation (9.1% vs. 10.4%, p=1.000), reoperation (10.4% vs. 13.0%, p=0.803), Type 1 neurological injury (11.7% vs. 13.0%, p=1.000), and ICU readmission (11.7% vs. 13.0%, p=1.000), but these differences were not statistically significant. Thirty-day mortality was also numerically lower in Group D but not statistically significant (11.7% vs. 14.3%, p=0.811).

**Conclusion:** Dexmedetomidine use during valvular cardiac surgery was associated with improved intraoperative hemodynamic stability, particularly through attenuation of heart rate responses at key surgical stress points. Postoperative complications including atrial fibrillation, acute kidney injury, neurological injury, reintubation, reoperation, ICU readmission, ICU length of stay, time to extubation, and 30-day mortality were numerically lower in the dexmedetomidine group. However these differences were not statistically significant. These findings suggest that dexmedetomidine may provide intraoperative hemodynamic benefits in patients undergoing valvular cardiac surgery, while its potential effects on postoperative organ protection and recovery outcomes require confirmation in larger multicenter randomized controlled trials.

## Introduction

Valvular heart disease continues to be a major public health challenge particularly in low and middle-income countries (LMICs) in India where Rheumatic Heart Disease(RHD) contributes significantly to disease burden^(1)^. Globally millions undergo cardiac surgery annually and patients with valvular pathologies face higher perioperative risks than those undergoing coronary artery bypass grafting ^(2,3)^. The physiological stresses of cardiopulmonary bypass (CPB), including systemic inflammation, ischemia-reperfusion injury and resultant sympathetic activation exacerbate these risks ^(4)^. Hemodynamic fluctuations perioperatively, arrhythmias, renal impairment and neurological injury remain frequent complications despite advances in surgical and anesthetic techniques ^(5)^.

Dexmedetomidine a highly selective α2-adrenoceptor agonist has emerged as a valuable adjunct in anesthesia and intensive care. It provides cooperative sedation without respiratory depression and exerts sympatholytic, anxiolytic and analgesic effects ^(6)^. Importantly its pharmacological actions extend beyond sedation: it causes reduction in catecholamine release, stabilizes heart rate and blood pressure, improves myocardial oxygen balance and modulates inflammatory cytokines ^(6,7)^. These properties make dexmedetomidine particularly attractive in the setting of cardiac surgery where hemodynamic stability and organ protection are critical in comparison to other types of surgeries.

Multiple studies have explored the role of dexmedetomidine in perioperative care. Systematic reviews and meta-analyses have demonstrated reductions in perioperative tachycardia, arrhythmia, delirium, and ICU stay ^(8,9)^. Trials in coronary artery bypass grafting and valve surgery suggest improved outcomes with dexmedetomidine, though evidence remains diverse ^(10,11,12)^. Cohort studies focusing on valve surgery populations specifically report reduced complications and shorter CICU length of stay ^(13)^. However gaps persist regarding consistent benefits on mortality and the risk of adverse effects such as bradycardia or hypotension which are known effects of the drug ^(14)^.

Given the particular vulnerability of patients undergoing valvular cardiac surgery, further evaluation of dexmedetomidine’s impact in this specific subgroup is essential. As the benefits which are being offered by the drug are fascinating notwithstanding the supposed adverse effects of the medication. This retrospective study was therefore undertaken to assess the effects of perioperative dexmedetomidine on intraoperative hemodynamics, postoperative complications, and ICU stay in adult patients undergoing valvular replacement surgery at a tertiary care center.

## Methods

### Study Design and Ethics

This was a retrospective observational study conducted at the Department of Cardiac Anaesthesia, Seth GS Medical College and KEM Hospital, Mumbai. Data were collected from institutional records between January 2022 and June 2023. Ethical approval was obtained from the Institutional Ethics Committee.

### Study Population

**Inclusion criteria:** Adults aged 18–70 years undergoing valvular cardiac surgery under CPB. **Exclusion criteria:** Preoperative opioid use, conduction disorders, oxygen therapy prior to inclusion, LVEF <40%, BMI ≥35 kg/m², shock, recent myocardial ischemia, adrenal insufficiency/on long term corticosteroid therapy, long term non-invasive ventilation, antecedent or active drug addiction, contraindications to dexmedetomidine or other anesthetic drugs, severe hepatic insufficiency, neurological disease, pregnancy/parturient or feeding mothers, pre-existing cognitive dysfunction, Person deprived of liberty by an administrative or judicial decision or person placed under judicial protection / under guardianship or guardianship and Patient participating in another drug trial or having participated in another drug trial within 1 month before randomization

### Group Allocation

- **Group C (Control, n=77):** Standard anesthesia without dexmedetomidine.
- **Group D (Dexmedetomidine, n=77):** Received IV dexmedetomidine infusion (0.5 μg/kg/hr) after induction until the end of surgery.

### Anesthetic Management

The patients on arrival to the operation theatre were connected to the monitor and baseline parameters (Heart rate, Spo2, Non-invasive blood pressure, Entropy and Near-infrared Spectroscopy) were recorded. Premedication with Inj. midazolam 0.02 mg/kg iv, Inj. Fentanyl 5-8ug/kg iv was given. Pre-oxygenation with 100% oxygen done for 3mins following which Inj. Dexmedetomidine was started as IV infusion at 0.5 ug/kg/hr for group D. For patients included in group C everything apart from the dexmedetomidine infusion was carried out in a similar fashion, which included supplemental fentanyl boluses for both groups at times where there was a decrease in depth of anesthesia. Baseline neuromuscular monitoring baseline for Train of Four was obtained after induction carried out using Inj. Etomidate 0.3mg/kg IV followed by muscle relaxant Inj. Rocuronium 1mg/kg. Anaesthesia was maintained with intermittent Inj. Rocuronium 0.1-0.15mg/kg, an infusion of dexmedetomidine at the rate of 0.5 μg/Kg/hour for group D and isoflurane. The infusion of dexmedetomidine was discontinued at the end of surgery.

### Cardiopulmonary Bypass and Monitoring

All patients underwent CPB using standard institutional protocols with moderate hypothermia (∼28 °C). CPB flow rates were maintained at 2.4 L/min/m², with mean arterial pressure (MAP) targeted at 60–70 mmHg. Arterial blood gases were monitored hourly. Hemodynamic monitoring included invasive arterial pressure, central venous pressure (CVP), entropy, and near-infrared spectroscopy (NIRS) for cerebral oxygenation. Heart rate (HR), MAP, CVP, entropy, and NIRS were recorded at predefined time points: baseline, post-induction, sternotomy, pericardiotomy, 5 minutes after protamine administration, and at the end of surgery.

### Postoperative Care

All patients were managed in the cardiac surgical ICU. Extubation, inotrope use and reintubation were guided by standard institutional protocols. Postoperative outcomes assessed included time of extubation, reintubation, reoperation, atrial fibrillation (AF), acute kidney injury (AKI, KDIGO criteria), type 1 neurological injury (stroke/TIA), ICU readmission, ICU length of stay (LOS), and 30-day mortality.

### Outcomes

- **Primary outcomes:** Intraoperative hemodynamic stability and entropy.
- **Secondary outcomes:** Postoperative complications (AF, AKI, reintubation, reoperation, neurological injury, mortality) and ICU LOS.

### Statistical Analysis

Continuous variables were expressed as mean ± SD and compared using Student’s t-test. Categorical variables were expressed as frequencies and percentages and compared using Chi-square or Fisher’s exact test, as appropriate. *P* <0.05 was considered statistically significant.

Analyses were performed using SPSS v22.

## Results

### Baseline Characteristics

Baseline demographics were comparable between groups.

### Intraoperative Hemodynamics

Group D demonstrated significantly lower HR at post-induction, sternotomy, and pericardiotomy compared with Group C (Table 2, Figure 1). MAP values were comparable between groups at baseline, post-induction, sternotomy, pericardiotomy, and end of surgery; however, MAP was significantly higher in Group D at 5 minutes after protamine administration (Table 2, Figure 2). CVP was significantly lower in Group D at baseline and at the end of surgery, while values at other intraoperative time points were comparable. NIRS and entropy values were not significantly different between groups across the measured time points (Table 2).

**Figure 1.**
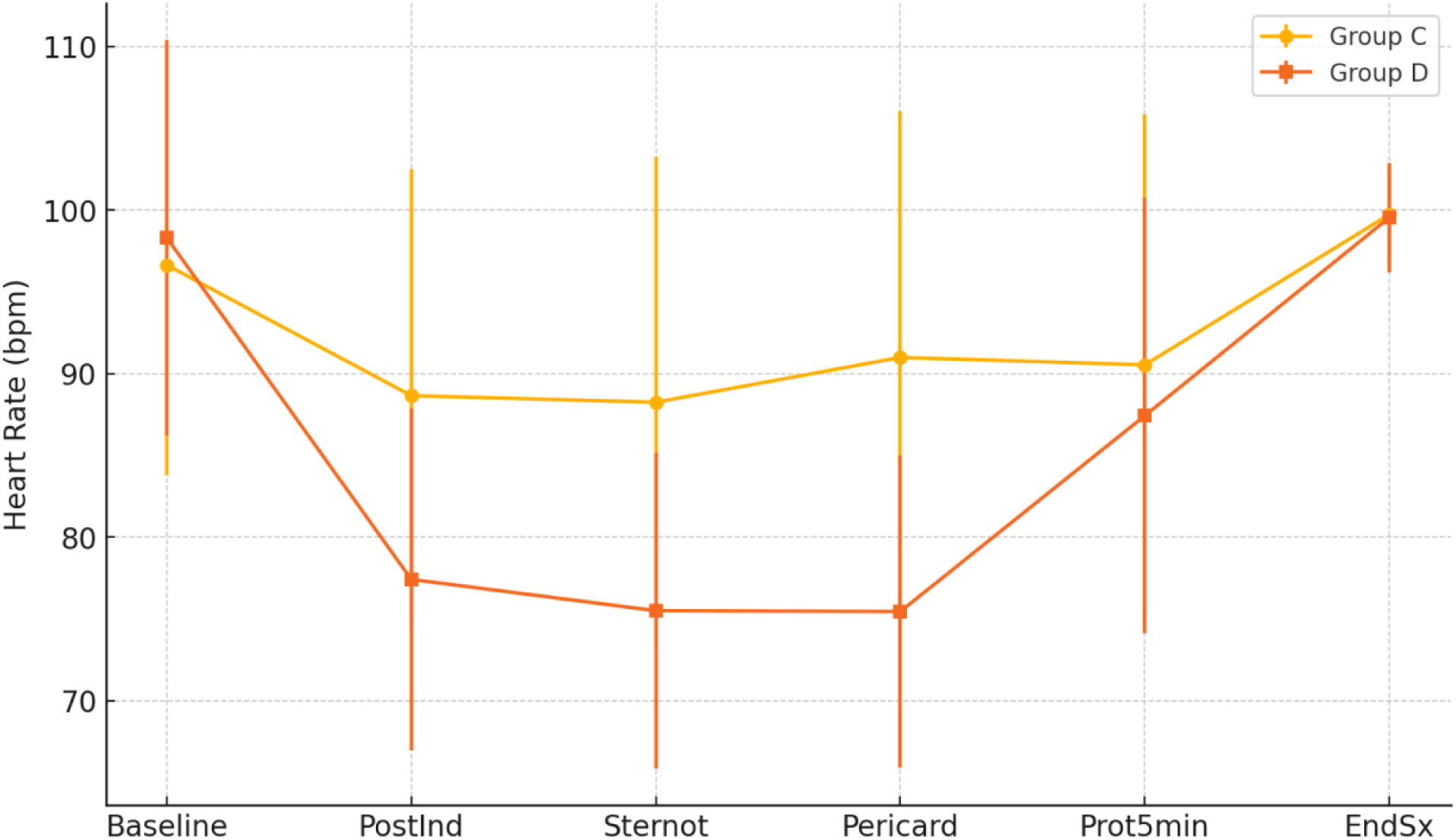
Intraoperative HR (mean ± SD) at six predefined time points.

**Figure 2.**
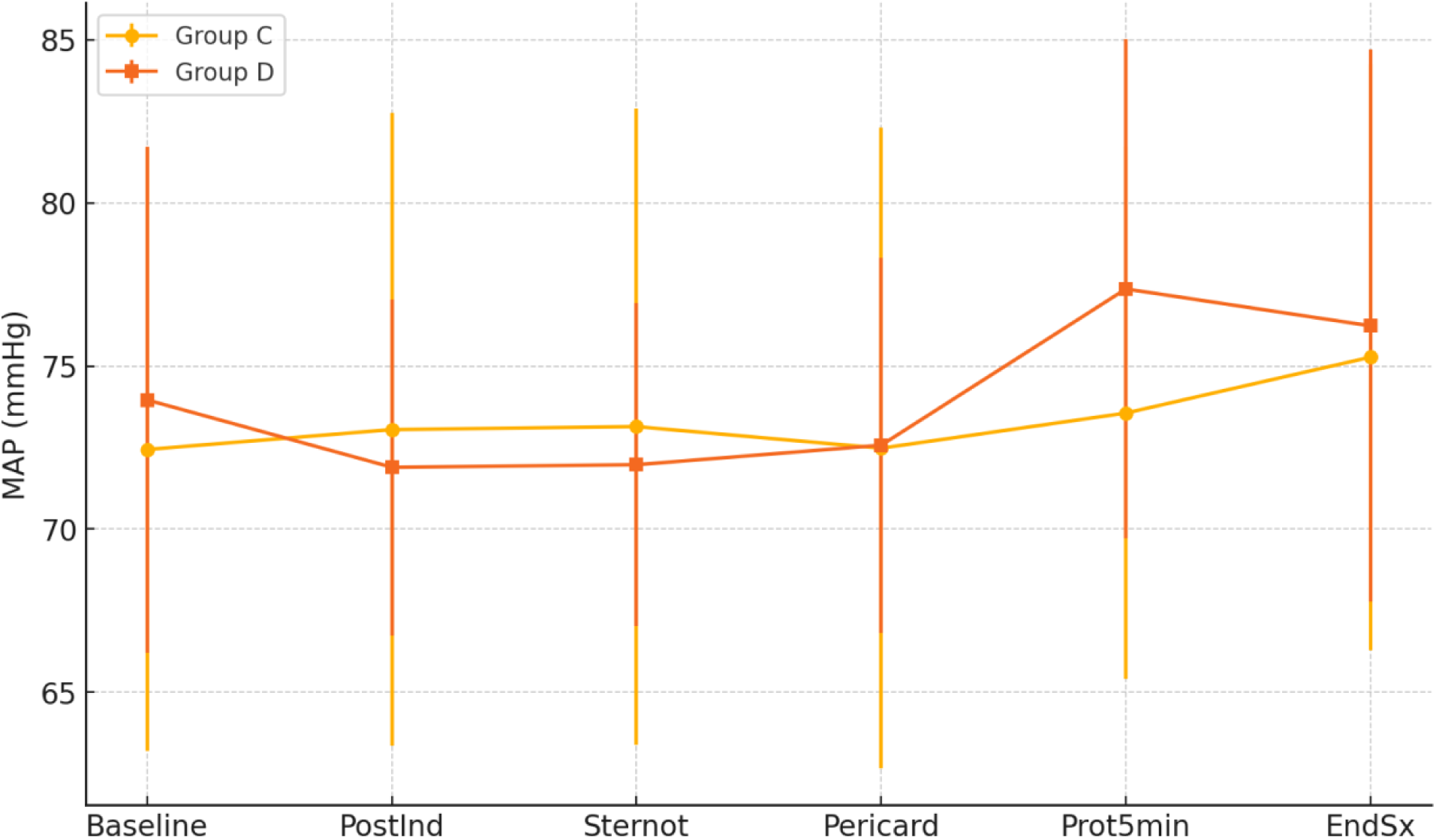
Intraoperative MAP (mean ± SD) at six predefined time points.

**Table 1.** Baseline Demographic and Clinical Characteristics.

| Variable | Group C (n=77) | Group D (n=77) | P value |
| --- | --- | --- | --- |
| Age (years) | 47.06 ± 13.88 | 43.13 ± 15.19 | 0.095 |
| ASA grade | 2.21 ± 0.77 | 2.00 ± 0.74 | 0.090 |
| Sex (F) | 38 (49.4%) | 35 (45.5%) | 0.747 |
| Sex (M) | 39 (50.6%) | 42 (54.5%) | 0.747 |

**Table 2.**
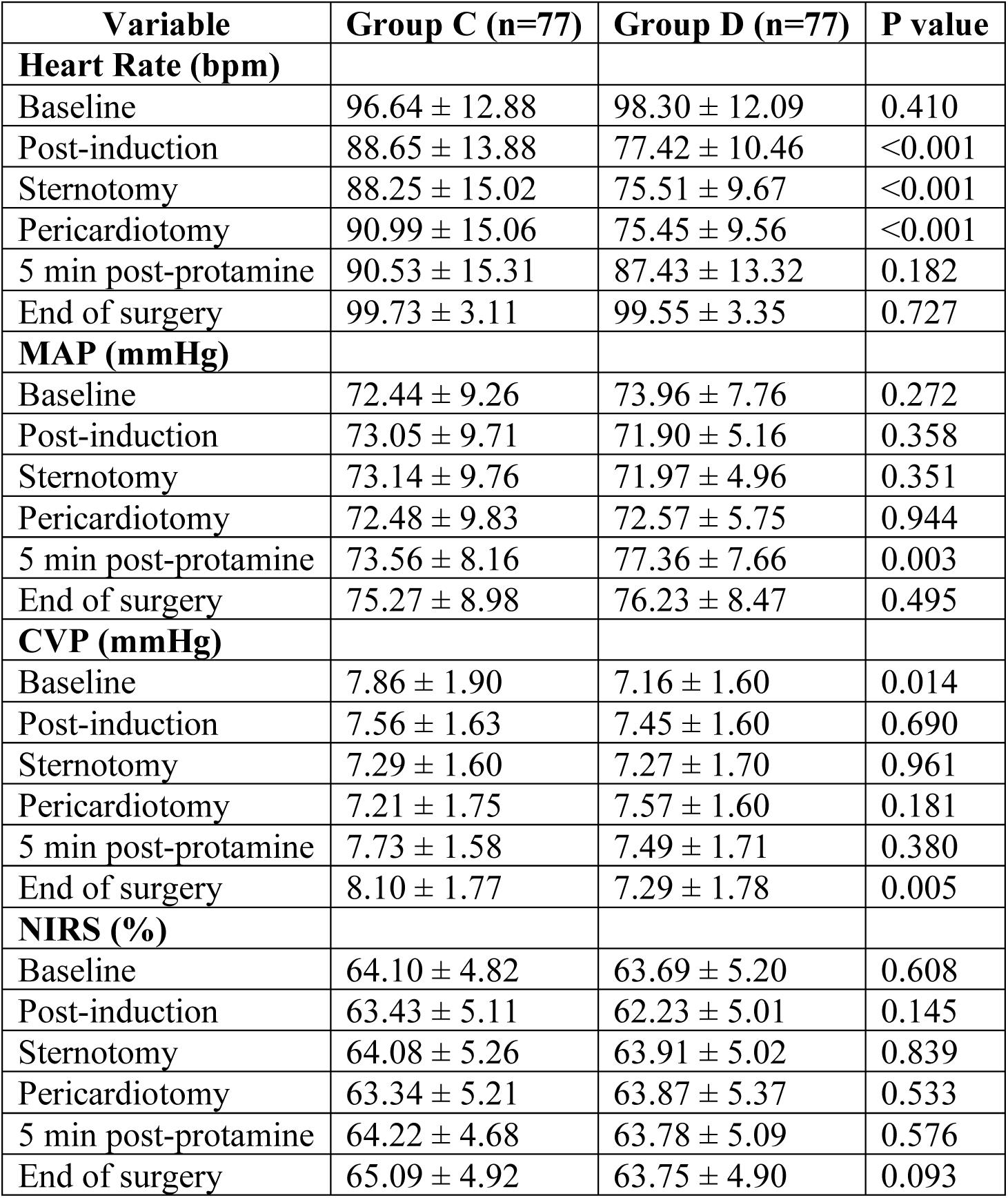
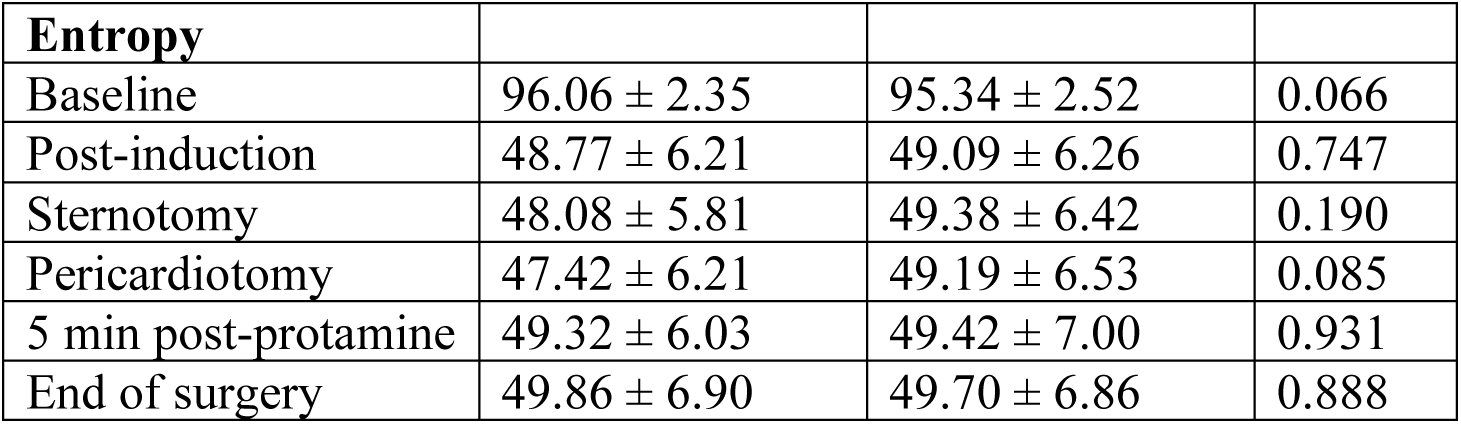
Intraoperative Hemodynamic Parameters.

### Postoperative Outcomes

Group D patients had numerically shorter ICU LOS and time to extubation compared with Group C; however, these differences were not statistically significant (Table 3, Figure 3). Postoperative complications, including atrial fibrillation, acute kidney injury, reintubation, reoperation, Type 1 neurological injury, ICU readmission, and 30-day mortality, were also numerically lower in Group D, but none of these differences reached statistical significance (Table 3, Figure 4A and 4B).

**Figure 3.**
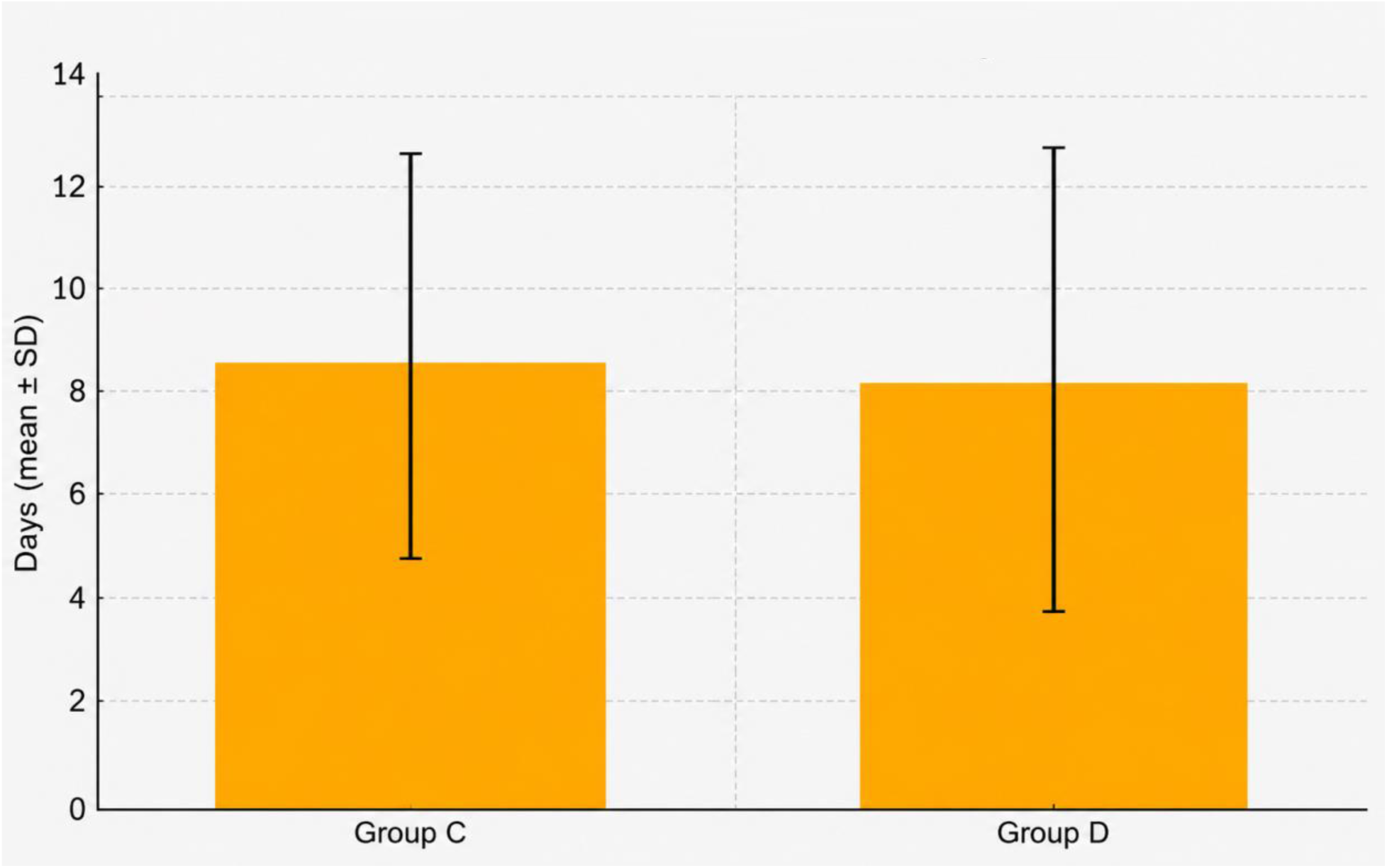
ICU length of stay (mean ± SD) in Group C vs Group D.

**Figure 4.**
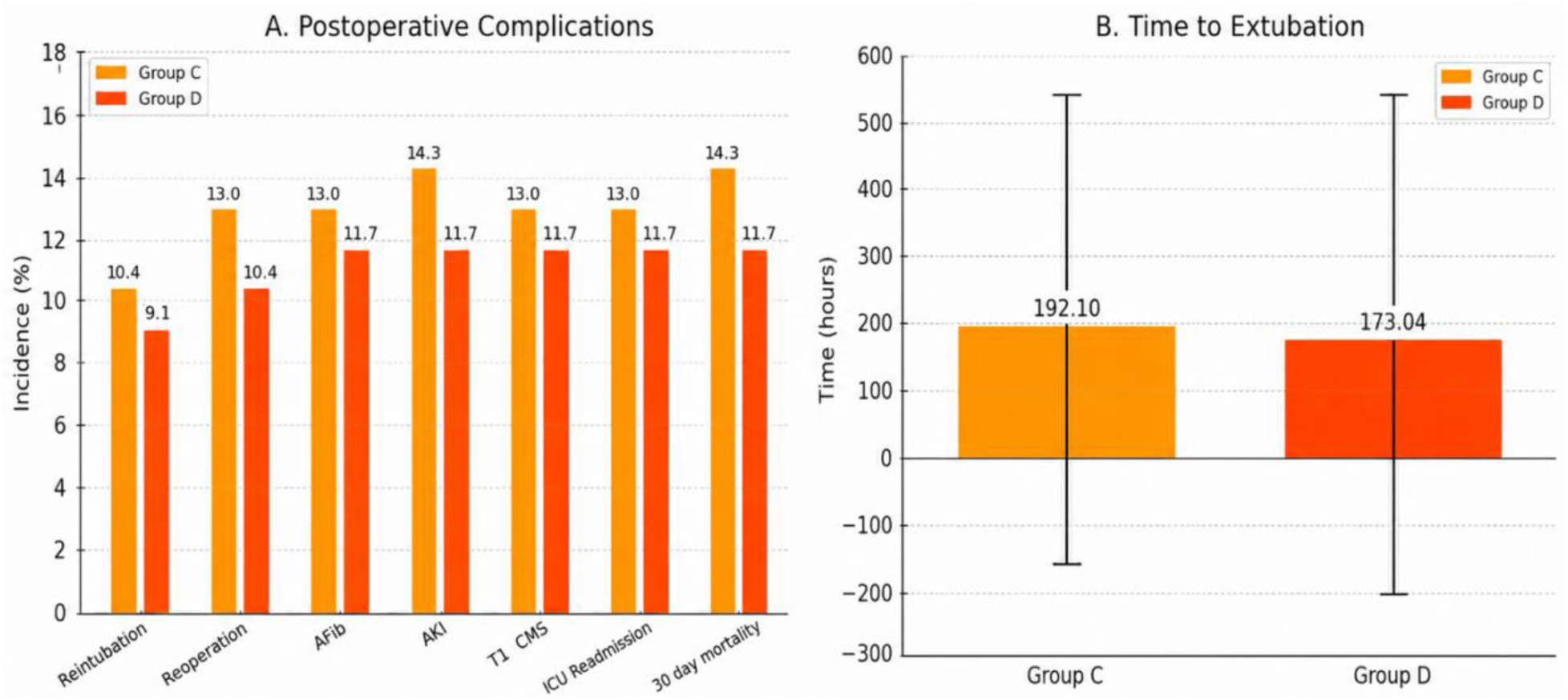
Postoperative outcomes. (A) Incidence (%) of postoperative complications including reintubation, reoperation, atrial fibrillation (AF), acute kidney injury (AKI), Type 1 neurological injury, ICU readmission, and 30-day mortality. (B) Time to extubation in Group C and Group D, expressed as mean ± SD in hours. Group D showed numerically lower postoperative complication rates and shorter time to extubation compared with Group C; however, these differences were not statistically significant.

**Table 3.**
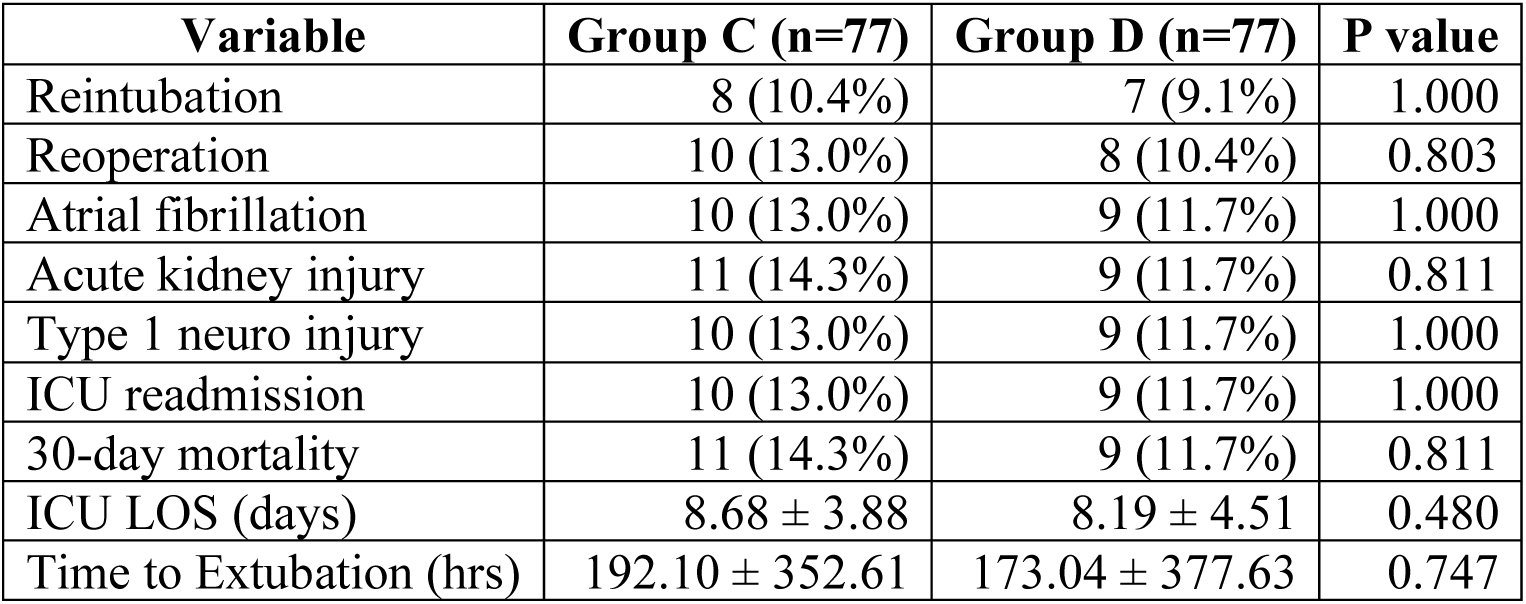
Postoperative Outcomes.

## Discussion

This study demonstrates that perioperative dexmedetomidine use in valvular cardiac surgery was associated with improved intraoperative hemodynamic stability, particularly through attenuation of heart rate responses at key surgical stress points. Although postoperative complications, ICU length of stay, time to extubation, and 30-day mortality were numerically lower in the dexmedetomidine group, these differences did not reach statistical significance in the revised dataset.

### Hemodynamic stability

Dexmedetomidine significantly lowered HR at post-induction, sternotomy, and pericardiotomy compared with the control group (Figure 1), consistent with its sympatholytic properties(5). MAP values were comparable between groups at most intraoperative time points, although Group D showed a significantly higher MAP at 5 minutes after protamine administration (Figure 2). These findings suggest that dexmedetomidine may attenuate perioperative tachycardic responses without causing clinically significant hypotension in this cohort, aligning with prior studies showing modulation of perioperative hemodynamic responses(7,8).

### Organ protection

Group D had numerically lower rates of AF, AKI, Type 1 neurological injury, reintubation, reoperation, ICU readmission, and 30-day mortality (Figure 4A). However, none of these postoperative outcomes were statistically significant in the revised analysis.

Therefore, while the observed trend is consistent with previously reported antiarrhythmic(12), renoprotective(9,15), and potential neuroprotective effects of dexmedetomidine(7,16,18), the present study cannot establish a definitive organ-protective effect. These findings should be interpreted as hypothesis-generating rather than confirmatory.

### Recovery

ICU LOS and time to extubation were numerically lower in Group D (Figures 3 and 4B), but the differences were not statistically significant. The lack of statistical significance may reflect the retrospective design, modest sample size, and wide variability in postoperative recovery times. Although dexmedetomidine has been associated with lighter cooperative sedation, opioid-sparing effects, and earlier recovery in previous studies(6,11), the revised findings from this cohort do not demonstrate a significant reduction in ICU stay or extubation time.

### Comparison with literature

Our intraoperative findings are consistent with prior literature describing dexmedetomidine-related attenuation of sympathetic responses during cardiac surgery. However, the revised postoperative results are more conservative than the earlier analysis and show only numerical, non-significant reductions in complications and recovery outcomes. This partially aligns with published studies and meta-analyses in which dexmedetomidine has shown variable effects on postoperative AF, AKI, ICU length of stay, mechanical ventilation duration, and mortality(8,10,13,14). The absence of statistically significant postoperative benefit in the present study may be related to limited sample size, event rates, and the single-center retrospective design.

### Strengths & limitations

Strengths include a homogenous valvular surgery cohort, comprehensive intraoperative monitoring, and analysis of clinically relevant postoperative outcomes. Limitations include the retrospective observational design, single-center scope, modest sample size, and the possibility of residual confounding. Additionally, the relatively low event rates and wide variability in recovery parameters may have limited the ability to detect statistically significant differences in postoperative outcomes.

### Clinical implications

Dexmedetomidine may be considered as a useful perioperative adjunct for improving intraoperative heart rate control in patients undergoing valvular cardiac surgery. However, based on the revised dataset, its role in reducing postoperative complications, shortening ICU stay, accelerating extubation, or lowering mortality remains uncertain. Larger prospective multicenter randomized controlled trials are required to clarify whether the numerical postoperative benefits observed in this study translate into clinically and statistically significant outcome improvements.

## Conclusion

Dexmedetomidine use during valvular cardiac surgery was associated with improved intraoperative hemodynamic stability, particularly through attenuation of heart rate responses at key surgical stress points. Postoperative complications including atrial fibrillation, acute kidney injury, neurological injury, reintubation, reoperation, ICU readmission, ICU length of stay, time to extubation, and 30-day mortality were numerically lower in the dexmedetomidine group; however, these differences were not statistically significant. These findings suggest that dexmedetomidine may provide intraoperative hemodynamic benefits in patients undergoing valvular cardiac surgery, while its potential role in postoperative organ protection and recovery outcomes requires confirmation in larger multicenter randomized controlled trials.

## Declarations

### Ethics Approval

This study was approved by the Institutional Ethics Committee of Seth GS Medical College and KEM Hospital, Mumbai (Project No:EC/OA-46-2024)

### Data Availability

The anonymized dataset is available from the corresponding author on reasonable request.

## Data Availability

All data produced in the present study are available upon reasonable request to the authors

## Acknowledgements

The authors thank the staff of the Department of Cardiac Anaesthesia, Seth GS Medical College and KEM Hospital, Mumbai, for their support in data collection

## Ethics Statement

This study was approved by the Institutional Ethics Committee of Seth GS Medical College and KEM Hospital, Mumbai (Project No: EC/OA-46/2024) The requirement for individual patient consent was waived due to the retrospective nature of the study.

## Author Contributions

- **Dr. Sanjeeta Umbarkar:** Conceptualization, methodology, manuscript review, critical revision
- **Dr. Jalaram Harshappan:** Data collection, Data interpretation, statistical analysis, manuscript drafting.
- **Dr. Parimal Pimpalkhute, Dr. Pooja Pimpalkhute, Dr. Renu Upadhyay, Dr. Juhi Kacha:** Approved the final manuscript.

## Data Availability Statement

The anonymized dataset analyzed during the current study is available from the corresponding author on reasonable request, in accordance with institutional and ethical guidelines.

